# Respiratory Infection Burden and Work Attendance among Healthcare Workers; The CHILL Study (Common Cold Healthcare Workers Immunological Longitudinal Learning)

**DOI:** 10.64898/2026.02.18.26346598

**Authors:** Mayan Gilboa, Noam Barda, Yael Weiss-Ottolenghi, Michal Canetti, Yovel Peretz, Ili Margalit, Gili Joseph, Michal Mandelboim, Yaniv Lustig, Gili Regev-Yochay

## Abstract

**Objective:** To quantify the seasonal burden of acute respiratory viral infections among healthcare workers (HCWs), characterize virologic etiologies, and identify predictors of symptomatic illness and sick leave.

**Methods:** We conducted a prospective cohort study of HCWs during winter 2024–2025, with weekly surveys capturing acute respiratory infections (ARI) and sick leave. Nasal–throat multiplex PCR swabs were self-collected during symptomatic episodes. Incidence rate ratios (IRRs) for symptomatic episodes and sick days were estimated using Poisson regression; presenteeism was assessed among febrile episodes.

**Results:** Among 655 HCWs, 400 (61.1%) reported ≥1 symptomatic episode. Over 70,861 person-days, incidence rates were 1.34 symptomatic episodes and 0.82 sick days per 100 person-days. Among PCR-confirmed episodes (n=112), rhinovirus (45.5%) and influenza (23.2%) predominated. Female sex was associated with higher rates of symptomatic episodes (IRR 1.38, 95% CI 1.11–1.72) and sick days (IRR 2.55, 95% CI 1.62–4.00), while age >56 years was associated with lower rates of both. During febrile episodes, 38.8% (95% CI 31.5%–46.6%) reported working despite fever.

**Conclusions:** ARIs were common among HCWs and frequently resulted in sick leave, yet febrile presenteeism remained substantial, underscoring the need for strengthened respiratory virus prevention and occupational health policies.

## Introduction

Acute respiratory infections (ARIs), predominantly caused by respiratory viruses, are a major, recurrent cause of morbidity and productivity loss, driving substantial healthcare utilization and work absenteeism (1,2). In the United States alone, ARIs account for ∼500 million episodes annually and an estimated ∼$40 billion in total costs (1). Although surveillance has traditionally focused on influenza, and more recently on SARS-CoV-2, other pathogens, including rhinovirus, respiratory syncytial virus (RSV), adenovirus, parainfluenza viruses, and human metapneumovirus (hMPV), circulate each winter and contribute to seasonal burden (3,4). Because clinical manifestations substantially overlap across etiologies, symptom-based case definitions have limited discriminatory value, emphasizing the need for virologic confirmation (5,6).

Existing surveillance systems, which largely derive from outpatient and hospital testing, preferentially capture more severe disease and therefore underestimate community incidence (7,8). Population-based cohort data indicate that many infections, including influenza, are mild or asymptomatic, and only a minority prompt medical attention (9). Among generally healthy adults, healthcare workers (HCWs) represent a priority population: they experience frequent exposures, have documented infection risk, and can contribute to onward transmission in healthcare settings (10–12). Pre-pandemic HCW cohorts demonstrate appreciable seasonal infection rates; for example, a multicenter study reported a 22.3% cumulative incidence of laboratory-confirmed influenza in a single season (13), with a substantial asymptomatic and pauci-symptomatic fraction that can still cause virus shedding and transmission (12,14).

Important knowledge gaps remain as most longitudinal data on ARIs were generated before the COVID-19 pandemic and focus almost exclusively on influenza(7,9,10). As a result, it remains uncertain how pandemic-era changes in viral circulation, healthcare-seeking behavior, and immunity from vaccination or prior infection have influenced ARI epidemiology among working adults, particularly healthcare workers. To address these gaps, we conducted a prospective cohort study with active follow-up and systematic PCR testing among HCWs at a tertiary medical center. Our objectives were to (i) quantify the frequency and distribution of symptomatic ARI episodes and identify the causative viruses; (ii) compare symptom profiles across pathogens; and (iii) characterize infection-linked symptomatic burden and sick-day utilization.

## Methods

### Study design, setting, and population

We conducted a prospective cohort study among healthcare workers (HCWs) at Sheba Medical Center, Israel, during the 2024–2025 winter season (October 1st 2024, to May 1st 2025). All staff aged ≥18 years were eligible. Enrollment was voluntary with written informed consent. Baseline demographics, occupational role, household characteristics, smoking history, and comorbidities were collected at enrollment (See survey in Supplementary Methods S1). The study was approved by the Sheba Medical Center institutional review board (IRB 0196-23).

### Weekly surveillance and outcome definitions

Participants completed weekly electronic questionnaires assessing acute respiratory symptoms and sick-day utilization (see questionnaire in Supplementary Methods S2). An acute respiratory infection (ARI) was defined as reporting ≥1 acute respiratory symptom on a weekly questionnaire. Fever episodes were symptomatic episodes with self-reported fever. Sick days were defined as the total number of self-reported days absent from work due to ARI during follow-up (see description of weekly surveillance and definition of episodes in Supplementary Methods S2). For the primary analysis, person-time was based on returned questionnaires; weeks without questionnaires were not counted toward follow-up, whereas completed questionnaires reporting no symptoms were considered asymptomatic weeks. To assess potential bias from weeks without returned questionnaires, we conducted a sensitivity analysis that incorporated non-response (described below).

### Specimen collection and laboratory testing

Episodes were eligible for virologic sampling if symptom onset occurred within the preceding 7 days. Eligible participants were instructed to self-collect combined nasal–throat swabs using standardized kits after 48 hours of symptoms, if symptoms persisted for 48 hours or more. Samples were delivered the same day when feasible or stored at −20°C until delivery. Multiplex real-time PCR testing was performed for influenza A/B, SARS-CoV-2, rhinovirus, respiratory-syncytial virus (RSV), adenovirus, parainfluenza viruses, and human metapneumovirus (hMPV) (see further description of Specimen collection and laboratory testing in Supplementary Methods S3).

### Statistical analysis

In the primary analysis, survey-derived person-time was defined as 7 days per completed weekly questionnaire (as the questionnaire specifically instructs the participants to answer based on the preceding 7 days). Crude incidence rates for symptomatic episodes, fever episodes, and sick days were calculated per 100 person-days. To estimate the association between different covariates and symptomatic episodes, fever episodes, and total sick days, we fit Poisson regression models with log(person-time) as the offset. Covariates were prespecified: sex, sector (administrative/other vs physician; nurse vs physician), age tertiles (≤40 years [reference], 41–56 years, >56 years), children ≤10 years at home, high-risk employee status, any chronic diagnosis, and past smoking. The association measure was the exponent of the coefficient of each covariate, interpreted as the incidence rate ratio.

All incidence rates are reported per person-day unless otherwise specified; weekly questionnaire-based analyses assume 7 person-days per completed questionnaire.

#### Virus-specific sick-leave burden

In a supplementary descriptive analysis, sick-leave burden was assessed at the episode level among PCR-confirmed symptomatic ARI episodes. Sick-leave days per episode were defined as zero when no sick leave was reported and as the recorded number of days when sick leave was reported. Analyses were restricted to single-virus detections with non-missing sick-leave data, and virus-specific burden was summarized using medians and interquartile ranges, as well as the proportion of episodes associated with any sick leave

**Presenteeism** was evaluated in episode-level models restricted to febrile episodes, defining sick leave as any sick day reported during that episode, with the same covariates used in the symptomatic episode model mentioned above.

For the primary analysis we used complete-case analyses. To evaluate potential bias from informative non-response at the weekly level, we constructed a person–week dataset from the first Monday on or after each participant’s enrollment date through 21 April 2025. For each participant–week, we defined questionnaire completion (response) based on the presence of a weekly survey in the questionnaires database and recorded weekly symptom status and sick-leave days when available. We then modeled the probability of questionnaire completion using logistic regression with sex, age tertiles, sector, and week in study as predictors, and derived stabilized inverse probability weights as the ratio of the overall response probability to the individual predicted probability for that week. Poisson models for weekly symptomatic episodes and weekly sick-leave days were refitted using these weights, with log(7) person-days as the offset and robust standard errors clustered by participant.

## Results

### Study population and follow-up

A total of 655 HCWs enrolled and contributed 10,123 weekly questionnaires over a 30-week period, corresponding to 70,861 survey-derived person-days of follow-up (Table 1). Participants had a mean age of 48.8±15.5 years and were predominantly female (463/655, 70.7%). Most were administrative/other staff (376/655, 57.4%), followed by nurses (149/655, 22.7%) and physicians (130/655, 19.8%). Baseline characteristics, including age, gender, sector distribution, household composition, comorbidity prevalence, and follow-up intensity, are summarized in Table 1. The age, sex, and sector distribution of the cohort was similar to that of the overall hospital workforce (Table 1)

**Table 1.**
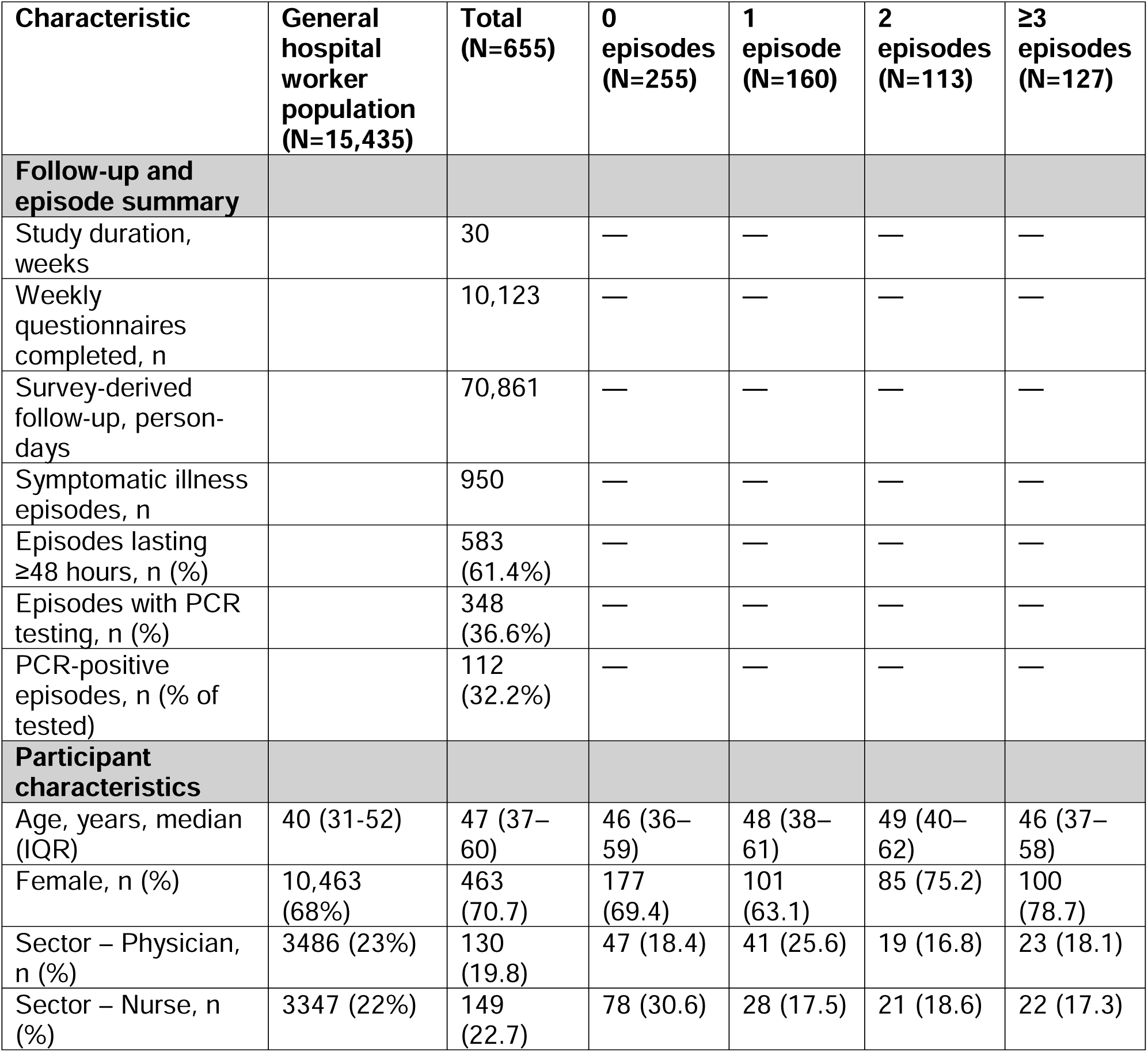
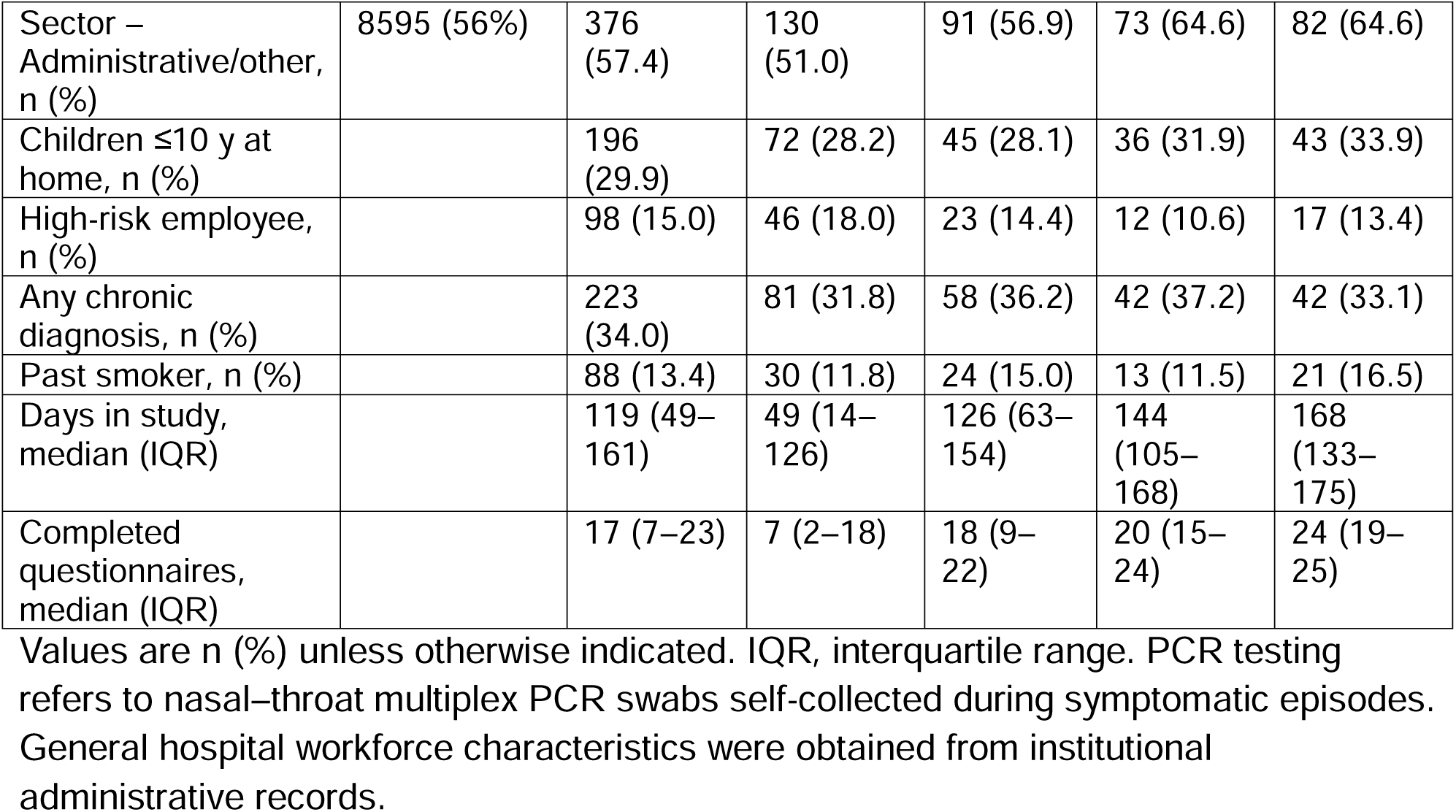
Characteristics of the study population by number of symptomatic respiratory episodes.

### Frequency, distribution, and timing of symptomatic ARI episodes

Respiratory illness was common: most participants (400/655; 61%) reported at least one symptomatic episode during the season, 37% (240/655) of the cohort reported recurrent illness, with a 127/655 (19%) experiencing three or more episodes (Table 1).Across the cohort, 950 symptomatic episodes were reported, the majority (583/950, 61%, 95% CI 58–65%) persisting >48 hours, and fever occurred in a minority of episodes (170/950, 18%, 95% CI 16–20%). Among the 626 participants who completed at least one weekly questionnaire, 70,861 survey-derived participant-days of follow-up were accrued, corresponding to crude incidences of 1.34 (95% CI 1.26–1.43) symptomatic episodes, 0.24 (95% CI 0.20–0.28) fever episodes, and 0.82 (95% CI 0.75–0.88) sick-leave days per 100 participant-days. Symptomatic activity exhibited clear seasonality, peaking in late December and January, mirroring national influenza-like illness trends reported through the Israeli Ministry of Health sentinel community surveillance (15) (Figure 1A).

**Figure 1.**
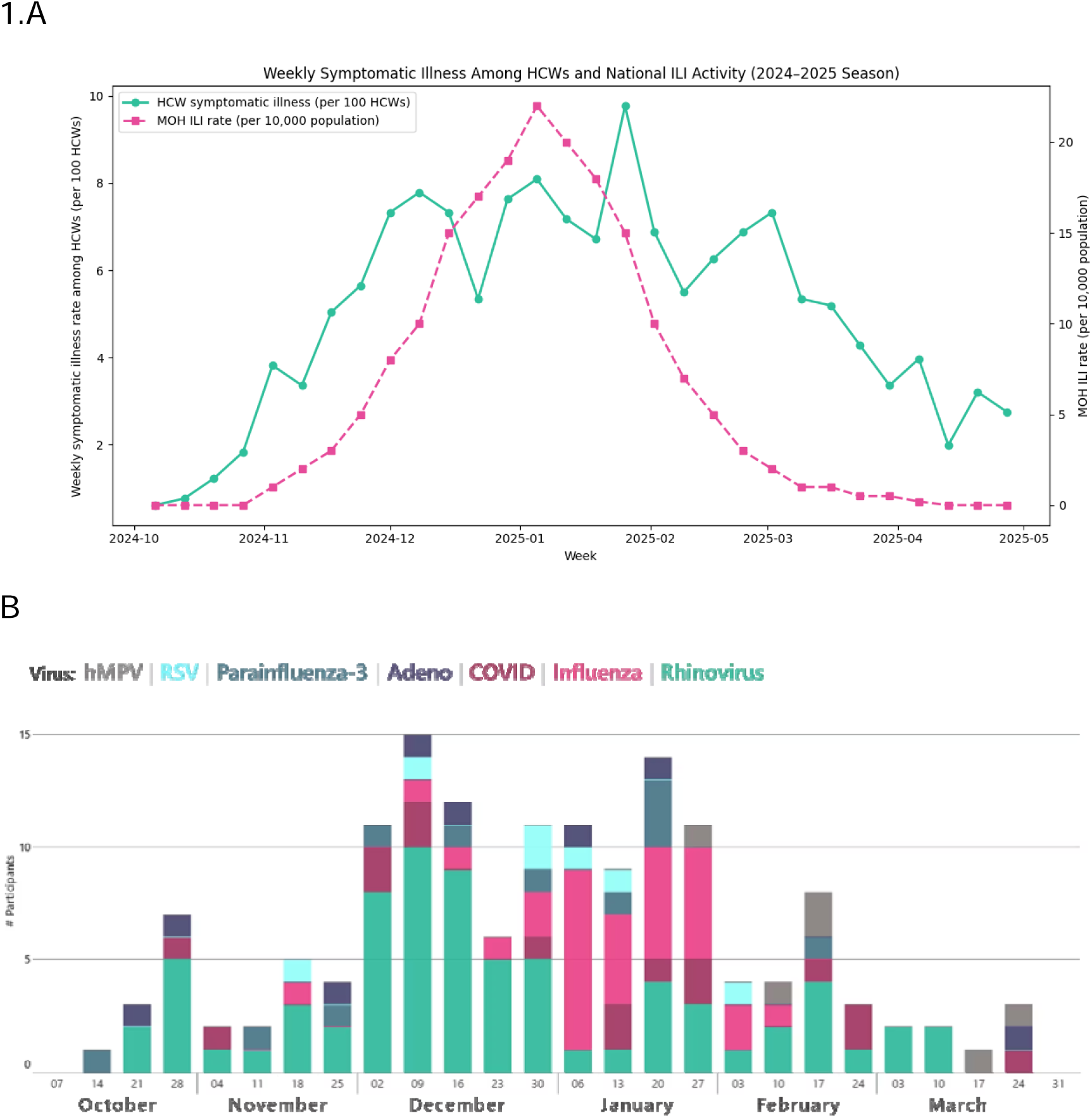
Seasonal timing of symptomatic respiratory illness and virologic detections in the CHILL cohort. A. Seasonal timing of symptomatic respiratory illness in the CHILL cohort and national influenza-like illness activity. Weekly incidence of self-reported symptomatic respiratory episodes in the CHILL cohort, expressed per 100 completed weekly questionnaires, over 30 weeks of surveillance (10,123 person-weeks), overlaid with contemporaneous national influenza-like illness (ILI) activity, expressed as cases per 10,000 population, as reported by the Ministry of Health (MOH). B. Respiratory virus detections over time among swabbed symptomatic episodes. Weekly distribution of respiratory virus detections identified by multiplex polymerase chain reaction (PCR) testing among symptomatic episodes with available virologic results (112 PCR-positive episodes out of 348 swabbed episodes). Bars represent the weekly counts of detected viruses **Abbreviations:** CHILL, Common cold Health care workers Immunological Longitudinal Learning; ILI, influenza-like illness; PCR, polymerase chain reaction; MOH, Ministry of Health.

In multivariable models for symptomatic episode rates, female sex was associated with higher symptomatic illness rates (IRR 1.38, 95% CI 1.11-1.72), whereas older age was associated with lower rates (age >56, compared to ≤40 years, IRR 0.72, 95% CI 0.55-0.94). Past smoking was associated with a modestly higher symptomatic episode rate (IRR 1.28, 95% CI 1.01-1.62). In contrast, living with children ≤10 years, high-risk ward employment, chronic diagnoses, and sector were not associated with symptomatic episode rates (Table 2A).

**Table 2.**
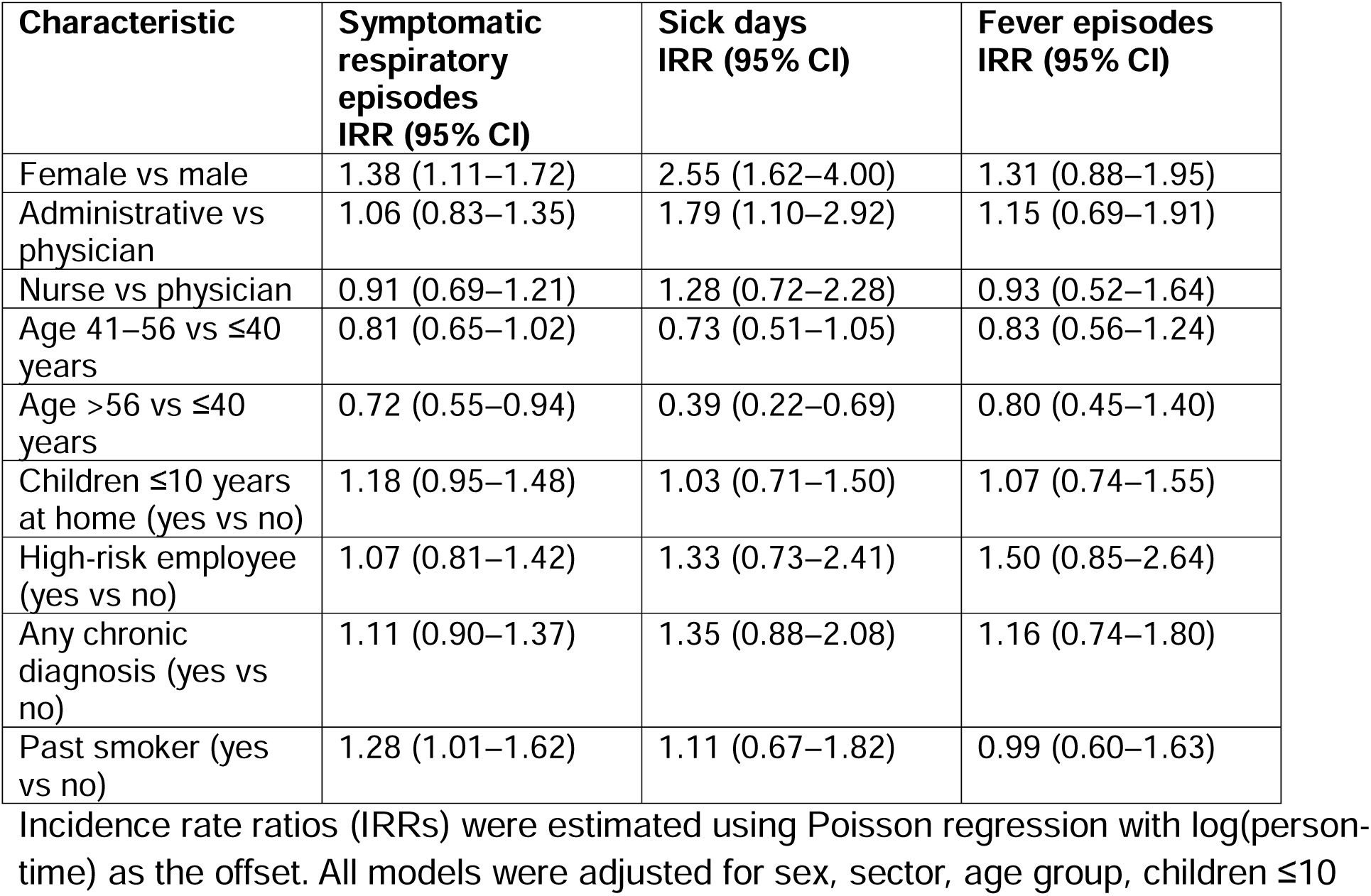
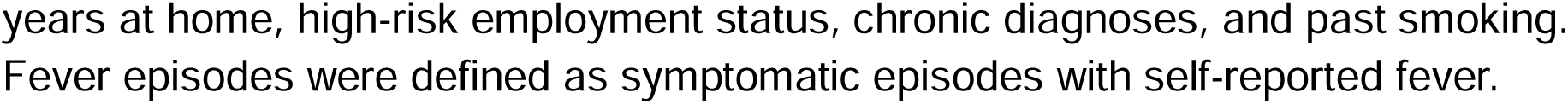
Association between participant characteristics and respiratory illness outcomes (primary analysis)

### Virologic etiologies

Among 950 symptomatic episodes lasting more than 48 hours, 348 (36.6%) underwent PCR testing; of these, 112 (32.2%) were positive for a respiratory virus. Rhinovirus was detected in 51/112 (45.5%), influenza A/B in 28/112 (25.0%), and SARS-CoV-2 in 11/112 (9.8%); parainfluenza virus in 9/112 (8.0%), human metapneumovirus (hMPV) in 6/112 (5.4%), respiratory syncytial virus (RSV) in 5/112 (4.5%), and adenovirus in 4/112 (3.6%). Co-infections were rare, occurring in 2/112 (1.8%) tested episodes (parainfluenza and influenza; influenza and adenovirus). The virologic time-course paralleled the clinical pattern: rhinovirus detections peaked in early December (10 positive episodes in a single week), whereas influenza detections peaked later, in the third week of January (5 positive episodes; Figure 1B).

### Symptom profiles across pathogens

Symptom distribution analyses were restricted to PCR-confirmed single-virus episodes with complete symptom data (n=89). Symptom patterns differed by virus (Figure 2). Among PCR-confirmed rhinovirus episodes (n=51), runny nose (36/51, 71%), sore throat (33/51, 65%) and cough (27/51, 53%) were most frequent, whereas fever was uncommon (6/51, 12%). In influenza episodes (n=28), systemic features were prominent: fever (17/28, 61%), fatigue (19/28, 68%), headache (18/28, 64%) and muscle pain (17/28, 61%) all occurred more often than in rhinovirus, alongside cough (19/28, 68%) and runny nose (21/28, 75%). SARS-CoV-2 episodes (n=11) generally resembled rhinovirus with more upper-respiratory syndromes: runny nose (7/11, 64%), cough (5/11, 45%) and sore throat (4/11, 36%) predominating, and fever reported only among one participant (1/11, 9%). Despite these differences in dominant features, there was substantial clinical overlap across pathogens.

**Figure 2.**
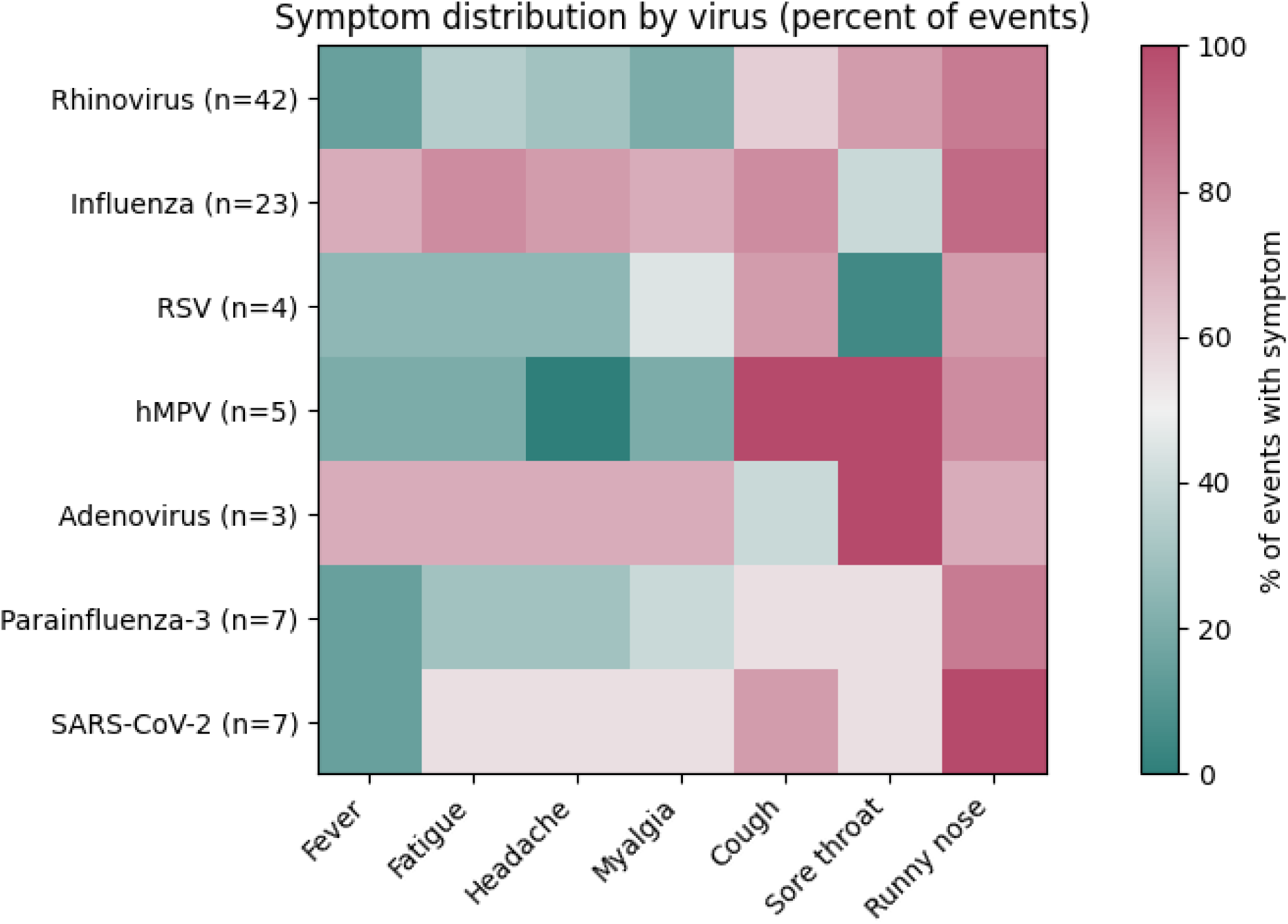
Symptom frequency by virus among PCR-confirmed symptomatic episodes. - Heatmap showing the proportion of PCR-confirmed episodes reporting each symptom, stratified by detected virus. Percentages are calculated within virus strata and among PCR-confirmed single-virus episodes with complete symptom data; denominators (n) are shown for each virus.

### Sick-day utilization and determinants of work absence

Sick leave was reported by 27% (179/655), of participants, with a median of 2 days per sick-leave episode. In adjusted models for sick days, female sex was associated with substantially higher sick-day rates (IRR 2.55, 95% CI 1.62-4.00), and administrative staff had higher sick-day rates than physicians (IRR 1.79, 95% CI 1.10-2.92). Older age was associated with fewer sick days (age >56, compared to ≤40 year, IRR 0.49, 95% CI 0.22-0.68). In contrast, children ≤10 years at home, high-risk employment, chronic diagnoses, and past smoking were not significantly associated with sick-day rates (Table 2B). Febrile episodes were less frequent than overall sick leave, occurring in 20% (131/655) of participants during follow-up. In adjusted models for febrile episodes, no demographic, household, or occupational characteristic was significantly associated with fever occurrence (Table 2C)

In inverse-probability–weighted weekly models that accounted for differential questionnaire completion, estimates were similar to the primary analysis. Women had higher weekly rates of symptomatic illness (IRR 1.36, 95% CI 1.10–1.69) and sick-leave days (IRR 2.45, 95% CI 1.55–3.88), administrative staff had higher sick-leave rates than physicians (IRR 1.75, 95% CI 1.08–2.83), and participants >56 years had markedly fewer sick-leave days than those ≤40 years (IRR 0.39, 95% CI 0.22–0.67) (supplementary table1 S1 and s2).

Among PCR-confirmed ARVI episodes with available sick-leave data, sick-leave burden differed by pathogen: influenza was associated with the greatest work-absence burden (median 1 day [IQR 0–2.5]; 56.5% with any sick leave), whereas rhinovirus and most other pathogens were associated with a median of 0 sick-leave days (Table S3)

### Presenteeism (episode-level analysis)

Among febrile episodes, 38.8% of participants reported working despite fever (66/170; 95% CI, 31.8%–46.2%). In participant-clustered episode-level models restricted to febrile episodes, we did not identify any significant associations between the prespecified covariates (sex, sector, age tertiles, children ≤10 years at home, high-risk employment, chronic diagnoses, or past smoking) and working despite fever. (Table 3).

**Table 3.** Factors associated with sick leave among symptomatic respiratory episodes (episode-level presenteeism analysis)

| Characteristic | Symptoms >48 h | Fever | Fever or symptoms >48 h |
| --- | --- | --- | --- |
| Episodes, n (% with sick leave) | 569 (204, 35.9%) | 170 (104, 61.2%) | 611 (225, 36.8%) |
| Female vs male, IRR (95% CI) | 1.72 (1.14–2.61) | 1.39 (0.92–2.09) | 1.70 (1.15–2.52) |
| Administrative vs physician, IRR (95% CI) | 1.06 (0.71–1.57) | 1.06 (0.66–1.70) | 1.11 (0.75–1.65) |
| Nurse vs physician, IRR (95% CI) | 1.00 (0.62–1.61) | 1.11 (0.65–1.87) | 1.04 (0.65–1.67) |
| Age 41–56 vs ≤40 years, IRR (95% CI) | 0.88 (0.68–1.15) | 1.00 (0.73–1.37) | 0.90 (0.69–1.16) |
| Age >56 vs ≤40 years, IRR (95% CI) | 0.57 (0.39–0.84) | 0.91 (0.62–1.34) | 0.60 (0.42–0.87) |

Analyses were restricted to symptomatic respiratory episodes. Sick leave was defined as ≥1 reported sick day during the episode. Symptoms >48 h indicates episodes lasting longer than 48 hours; Fever indicates episodes with self-reported fever; Fever or symptoms >48 h includes episodes meeting either criterion. Incidence rate ratios (IRRs) were estimated using participant-clustered Poisson regression models adjusted for sex, sector, age group, children ≤10 years at home, high-risk employment status, chronic diagnoses, and past smoking.

## Discussion

In this prospective cohort of 655 healthcare workers followed with weekly active surveillance across the 2024-2025 winter season, respiratory illness was frequent and operationally consequential: 61.1% reported at least one symptomatic episode, with substantial recurrence (37% reporting ≥2 episodes), and 578 sick days accrued. Among swabbed symptomatic episodes, rhinovirus and influenza predominated, while symptom profiles varied by pathogen they remained clinically overlapping. This finding is consistent with prior evidence that clinical features alone have limited ability to distinguish respiratory viral etiologies without virologic confirmation (16).

Three findings merit particular emphasis. First, the respiratory illness burden among HCWs was frequent and recurrent. Over the winter season, more than half of the participants experienced at least one symptomatic episode, with substantial recurrence. This burden far exceeds rates captured by conventional healthcare-based surveillance systems, which rely primarily on medically attended illness and laboratory-confirmed testing (17). The large gap between conventionally captured episodes and the burden reported in this study suggest that a substantial share of ARI burden is underrepresented in conventional surveillance data. This pattern is consistent with community- and HCW-based active surveillance studies showing that respiratory illnesses are common, often mild, and incompletely captured by healthcare utilization (9,18–20). In these cohorts, as in our study, rhinovirus predominates in incidence, whereas influenza, despite lower frequency, accounts for a disproportionate share of seasonal morbidity. The operational footprint of this burden is substantial. Extrapolating the observed crude sick-leave rate in our cohort to a ∼15,000-employee hospital implies a substantial wintertime loss of staff-days per day. This is consistent with hospital-based observations that respiratory virus seasons measurably increase healthcare-worker absenteeism and operational strain (21).

Second, we observed a consistent demographic gradient in both illness frequency and absenteeism. Female sex was independently associated with higher rates of symptomatic episodes and sick-day use, whereas older age was associated with less sick days. These gradients persisted after adjustment for plausible exposure proxies: including young children at home, employment in high-risk wards, and chronic diagnoses, which were not independently associated with either outcome. The lack of association with simple household composition does not exclude a role for household transmission and may, in part, reflect limited power to detect modest effects; however, it also suggests that such coarse measures may not capture relevant exposure pathways. Gendered differences in caregiving responsibilities, intensity and duration of child–adult contact, and decision-making around work attendance are unlikely to be captured in a binary children-at-home variable. Consistent with this interpretation, household transmission analyses from the Israeli COVID-19 Family Study demonstrated higher child-to–female adult transmission risk than child-to–male adult transmission(22).

Similar sex- and age-based gradients have been reported in other Israeli healthcare-worker cohorts during winter respiratory virus seasons (18), supporting the interpretation that these patterns reflect persistent structural and behavioral determinants rather than study-specific artifacts. Evidence from prospective influenza surveillance further highlights how symptom perception, reporting thresholds, and surveillance design shape observed illness burden (12,13). Together, these findings suggest that the gradients observed in CHILL reflect a combination of contact structure, reporting behavior, and practical constraints on taking sick leave.

Third, febrile presenteeism was common and directly relevant to infection prevention. Nearly two in five febrile episodes were not associated with sick leave, indicating that many HCWs arrive to work despite fever. This finding aligns with prior prospective studies showing that working while ill is common among HCWs and that viral shedding can occur during on-duty illness (12,23).

This study has several limitations. Participation was voluntary and included only a small subset of the institution’s work force (approximately 15,000 HCWs), which may introduce selection bias and limit representativeness. Although the demographic composition of the cohort was broadly similar to the overall healthcare workforce at the institution with respect to sex, age distribution, and occupational sector, individuals who choose to participate in intensive longitudinal surveillance studies may be more attentive to symptoms and more likely to report illness, potentially inflating observed illness frequency; participants may differ from non-participants in exposure patterns, health behaviors, and willingness to report illness or take sick leave. Questionnaire completion was incomplete, creating potential for informative non-response if missing surveys were related to symptoms or workload; we addressed this through sensitivity analyses using a missingness-pattern-informed weighting scheme, but residual bias cannot be excluded. Virologic testing was performed for only a subset of symptomatic episodes and likely reflected both symptom severity and participant compliance, which may bias pathogen distributions and limits etiologic inference, however, the temporal patterns of influenza observed in our cohort closely mirrored those captured by contemporaneous national surveillance, suggesting that, despite incomplete testing, the study was able to capture broader seasonal trends. Symptoms, fever, and sick leave were self-reported and may be subject to reporting and recall biases; however, surveys were administered weekly with additional electronic reminders, which likely reduced the impact of these biases. The relatively small number of febrile episodes reduced power for pathogen- and severity-stratified analyses. Finally, this single-center study reflects local policies and culture around illness and absenteeism, and findings may not generalize to other hospitals or seasons. However, Sheba Medical Center is a large tertiary academic hospital with a diverse workforce, participant demographics closely mirrored the overall staff composition, and similar seasonal and behavioral patterns have been reported in other healthcare worker cohorts, supporting the broader relevance of our findings(18,19).

In conclusion, active weekly surveillance demonstrates that winter ARI among HCWs is frequent, often recurrent, and operationally important at health-system scale, with substantial absenteeism and persistent febrile presenteeism. These data strengthen the case for institutional strategies that reduce barriers to appropriate sick leave and support safer work practices during symptomatic, and particularly febrile-illness, alongside broader surveillance approaches that capture true workforce burden beyond medically attended testing, and reinforce the importance of seasonal influenza and COVID-19 vaccination and consistent masking during respiratory virus season to mitigate transmission and preserve staffing capacity.

## Ethical approval

The study was approved by the Sheba Medical Center Institutional Review Board (IRB No. 0196-23). Written informed consent was obtained from all participants.

## Data Availability

All data produced in the present study are available upon reasonable request to the authors

## Acknowledgments

We thank Lilach Or, Shiraz Katovitz, Keren Tal, and Almog Cohen-Huszti for their important contributions to participant recruitment and longitudinal follow-up of the cohort. We are grateful to the Infection Prevention and Control Unit and the SPRI team for their essential assistance and support throughout the study. Finally, we sincerely thank all participating healthcare workers who dedicated time from their very busy schedules to advance scientific knowledge.

---

**Supplementary Table S1.** Weekly IPW-adjusted associations with symptomatic illness and sick-leave days.

| Characteristic | Symptomatic weeks<br>IRR (95% CI) | Sick-leave days<br>IRR (95% CI) |
| --- | --- | --- |
| Female vs male | 1.36 (1.10–1.69) | 2.45 (1.55–3.89) |
| Administrative vs physician | 1.14 (0.89–1.46) | 1.74 (1.08–2.82) |
| Nurse vs physician | 1.00 (0.77–1.31) | 1.27 (0.73–2.23) |
| Age 41–56 vs ≤40 years | 0.75 (0.61–0.93) | 0.72 (0.50–1.04) |
| Age >56 vs ≤40 years | 0.69 (0.54–0.88) | 0.39 (0.22–0.67) |
| Children ≤10 y at home<br>(yes vs no) | 1.23 (0.99–1.53) | 1.01 (0.70–1.46) |
| High-risk employee (yes vs<br>no) | 1.13 (0.87–1.48) | 1.28 (0.71–2.30) |
| Any chronic diagnosis (yes<br>vs no) | 1.11 (0.91–1.35) | 1.34 (0.88–2.03) |
| Past smoker (yes vs no) | 1.26 (1.00–1.58) | 1.13 (0.69–1.86) |

Incidence rate ratios (IRRs) were estimated using weekly Poisson regression models with a log link and robust standard errors clustered by participant. Inverse probability weights were derived from a logistic regression model for weekly questionnaire completion conditional on sex, age group, occupational sector, and week in study. An offset of log(7 person-days per week) was applied

**Supplementary Table S2.** Sick-leave burden per PCR-confirmed ARI episode, by virus.

| Virus (PCR-confirmed) | PCR+ episodes, N | Episodes with sick-leave data, n/N (%) | Any sick leave, n (%) | Median sick-leave days (IQR) | Mean (SD) |
| --- | --- | --- | --- | --- | --- |
| **Rhinovirus** | 51 | 41/51<br>(80.4%) | 10 (24.4%) | 0 (0–0) | 0.61 (1.20) |
| **Influenza** | 26 | 23/26<br>(88.5%) | 13 (56.5%) | 1 (0–2.5) | 1.61 (2.02) |
| **SARS-CoV-2** | 11 | 7/11<br>(63.6%) | 2 (28.6%) | 0 (0–1) | 0.71 (1.25) |
| **Parainfluenza**<br>**virus type 3** | 8 | 7/8<br>(87.5%) | 0 (0.0%) | 0 (0–0) | 0.00 (0.00) |
| **Human**<br>**metapneumovirus** | 6 | 5/6<br>(83.3%) | 1 (20.0%) | 0 (0–0) | 0.40 (0.89) |
| **RSV** | 5 | 4/5<br>(80.0%) | 1 (25.0%) | 0 (0–0.25) | 0.25 (0.50) |
| **Adenovirus** | 3 | 3/3<br>(100.0%) | 2 (66.7%) | 1 (0.5–1.5) | 1.00 (1.00) |
| **Overall** | 110 | 90/110<br>(81.8%) | 29 (32.2%) | 0 (0–1) | 0.81 (1.44) |

Sick-leave days per episode were defined as zero when no sick leave was reported and as the recorded number of days when sick leave was reported. Analyses were restricted to single-virus PCR-confirmed episodes with available sick-leave data.
Footnote: Co-detections (n=2) were excluded. Sick-leave data were missing for 20/110 (18.2%) single-virus PCR-confirmed episodes.

## Supplementary Methods S1. Enrollment and baseline variables

Participants provided written informed consent and completed a baseline questionnaire capturing age, sex, sector (physician/nurse/administrative-other), household composition, work location, smoking history, and comorbidities. **Children** ≤**10 years at home** was defined as living with children and youngest child aged ≤10 years. **High-risk employee** was defined as frontline work in emergency department, pediatrics, internal medicine, or intensive care units. **Any chronic diagnosis** was coded as present if any of the following were reported: hypertension, hyperlipidemia, diabetes, heart disease, chronic pulmonary disease, coagulation disorder, chronic liver disease, chronic kidney disease, autoimmune disease, immunosuppression. Smoking was categorized as never vs past; no participants reported current smoking.

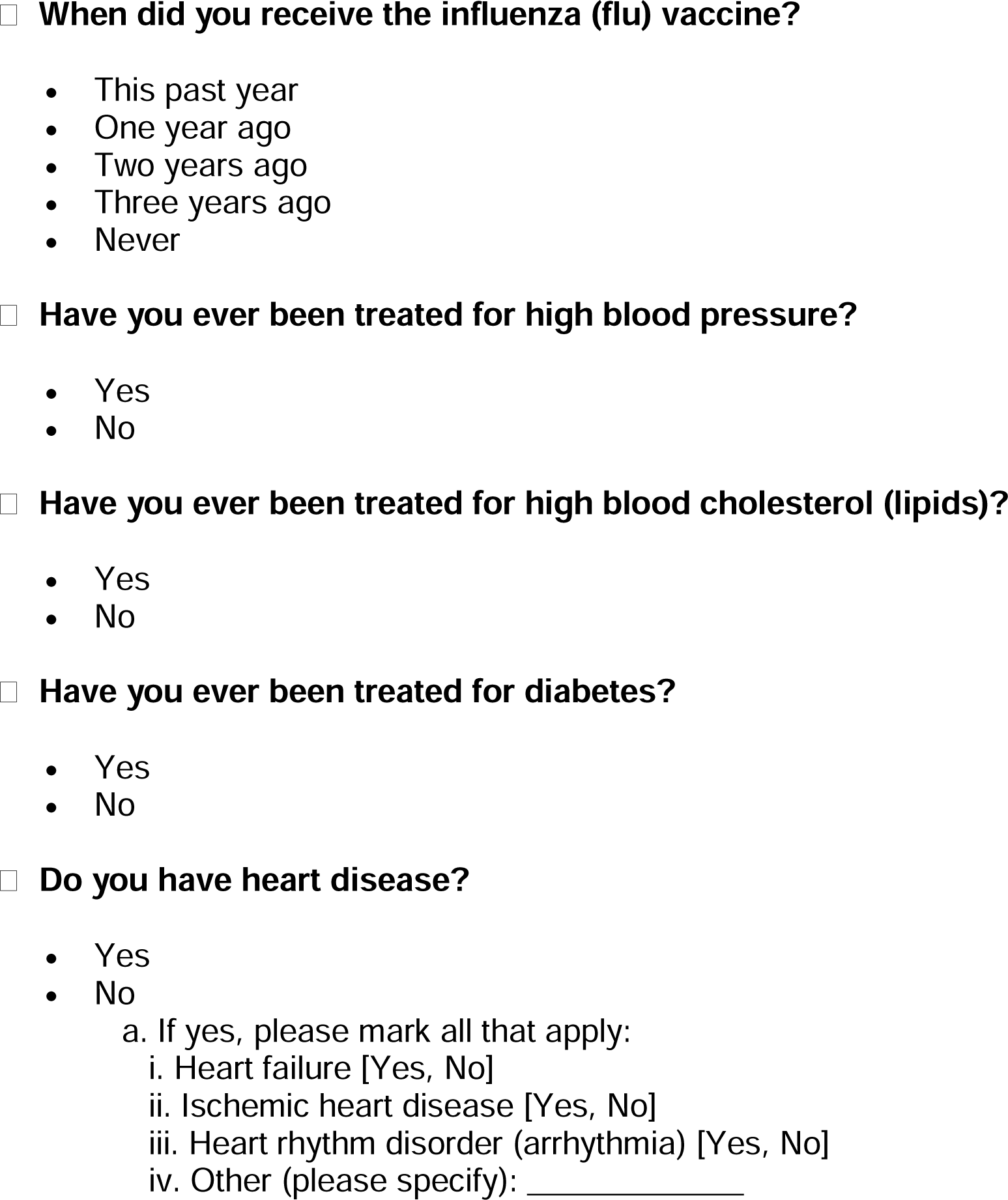
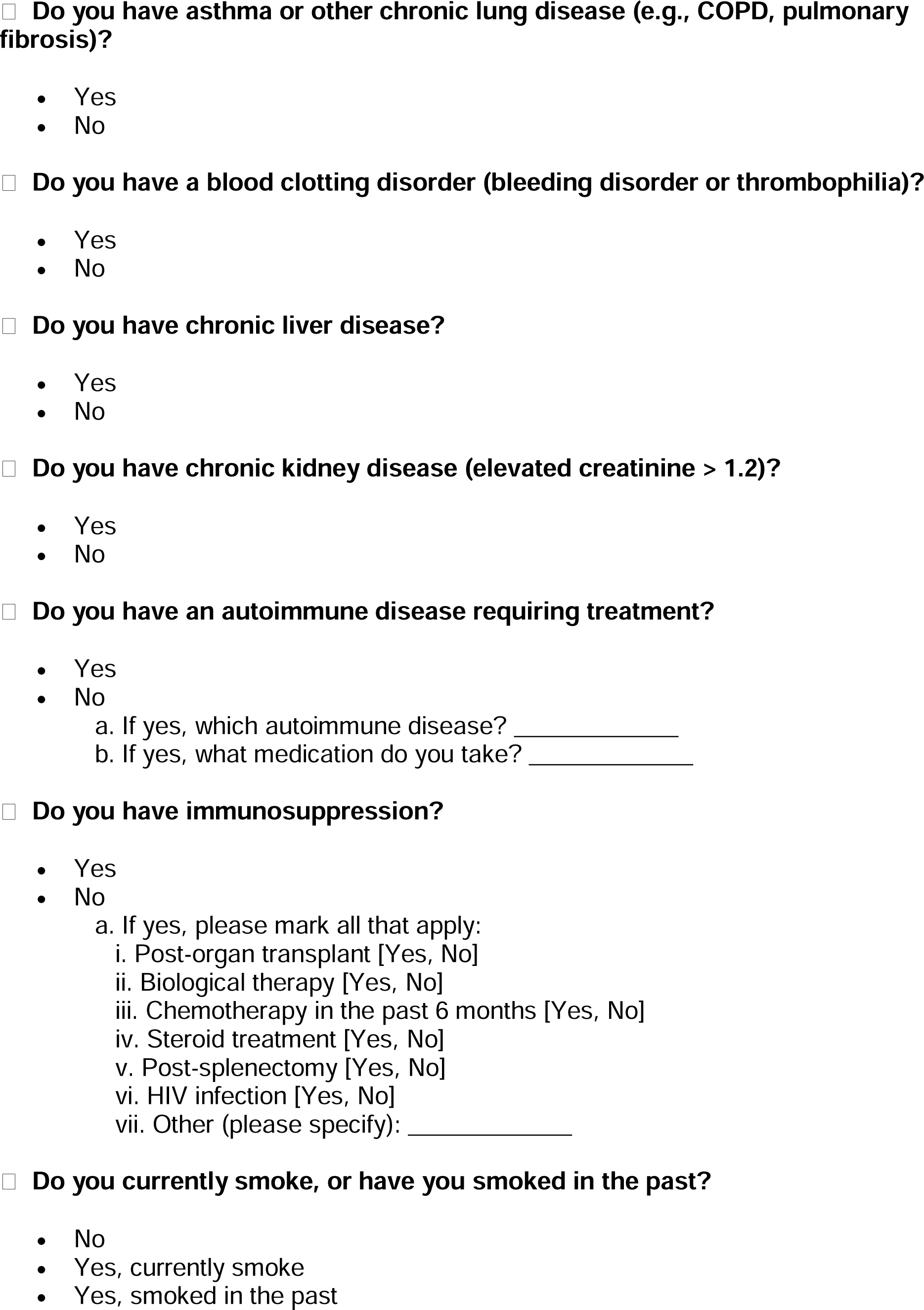
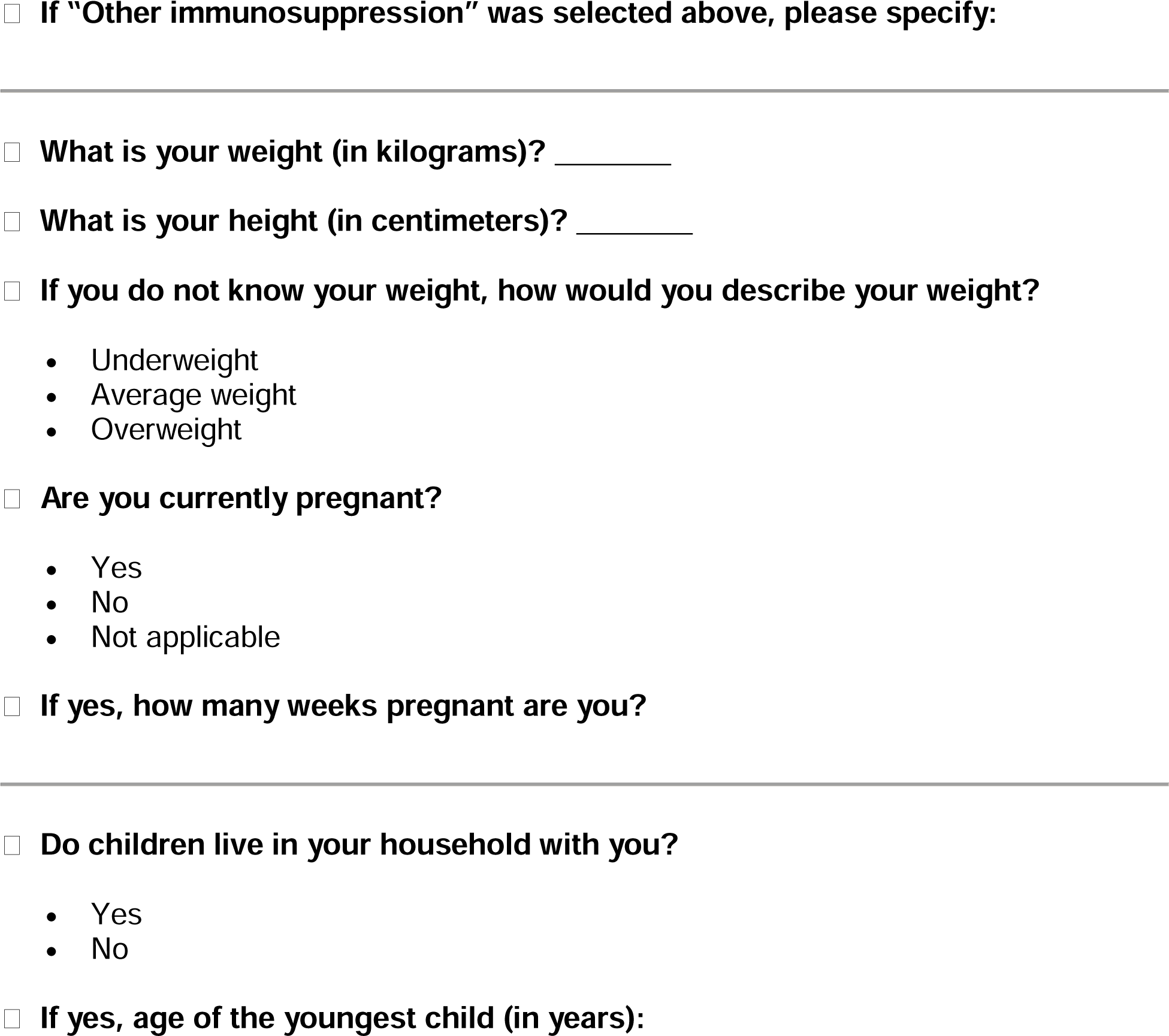

## Supplementary Methods S2. Weekly surveillance and episode construction

Weekly REDCap questionnaires captured: (i) presence/absence of acute respiratory symptoms (including fever, cough, sore throat, rhinorrhea, fatigue, myalgia); (ii) symptom duration including whether symptoms persisted >48 hours; and (iii) number of sick days in the preceding week. Participants were encouraged to report mild symptoms. A symptomatic episode was defined as ≥1 symptom in a weekly questionnaire. Symptoms >48 hours was defined as self-reported duration >48 hours within the symptomatic week. Episode-level sick leave was defined as any report of ≥1 sick day in that symptomatic week; total sick days was the sum across all completed questionnaires. For the primary analysis, only weeks with completed questionnaires contributed person-time; for the calendar-time sensitivity analysis, time accrued regardless of questionnaire completion.

Symptom survey:

1. Have you experienced any flu-like symptoms (fever, sore throat, headache, myalgia, etc.) during the past week? [Yes, No]
2. Did the symptoms last longer than 48 hours? [Yes, No]
3. Please mark which of the following symptoms you have:

a. Fever lasting up to 48 hours [Yes, No]
b. Fever lasting more than 48 hours [Yes, No]
c. Dyspnea [Yes, No]
d. Cough [Yes, No]
e. Myalgia [Yes, No]
f. Severe fatigue or weakness [Yes, No]
g. Loss of taste and/or smell [Yes, No]
h. Runny nose [Yes, No]
i. Headache [Yes, No]
j. Sore throat [Yes, No]
k. Abdominal pain/ nausea/ vomiting [Yes, No]
l. Other [please specify]
4. Did you require any medication? [Yes, No]
5. Have you been tested with a COVID-19 or influenza antigen test? [Yes, No]
6. Have you had COVID-19, the flu, or any other respiratory illness in the past year? [Yes, No]
7. Have you taken sick leave in the past week? [Yes, No]
8. If yes, how many?

## Supplementary Methods S3. Specimen collection and laboratory testing

Symptomatic episodes were eligible for sampling if reported symptom onset was within 7 days. Participants self-collected combined nasal–throat swabs using standardized kits and delivered samples the same day when possible; otherwise, samples were stored at −20°C until delivery. Nucleic acid extraction and multiplex real-time PCR were performed in the hospital virology laboratory for influenza A/B, SARS-CoV-2, rhinovirus, RSV, adenovirus, parainfluenza viruses, and hMPV. Co-infection was defined as detection of >1 virus in the same sample.

### Viral RNA extraction and real-time PCR

Viral RNA was extracted using the STARMag Viral DNA/RNA 200C universal kit (Seegene Inc., Korea). Reverse transcription and RT-PCR assays were performed using the Seegene Allplex™ RV Master assay is a one-step multiplex real-time RT-PCR system for detecting eight viral respiratory pathogens including; severe acute respiratory syndrome coronavirus 2 (SARS-CoV-2), Influenza A, Influenza B, Human respiratory syncytial virus (RSV), adenovirus, rhinovirus, parainfluenza virus (1–4), and metapneumovirus (MPV) (Seegene Inc., Korea). The assays were performed in CFX real-time PCR system (BIO-RAD, United States).

